# End-to-End Machine Learning based Discrimination of Neoplastic and Non-neoplastic Intracerebral Hemorrhage on Computed Tomography

**DOI:** 10.1101/2024.09.30.24314346

**Authors:** Jawed Nawabi, Sophia Schulze-Weddige, Georg Lukas Baumgärtner, Tobias Orth, Andrea Dell Orco, Andrea Morotti, Federico Mazzacane, Helge Kniep, Uta Hanning, Michael Scheel, Jens Fiehler, Tobias Penzkofer

**Author notes:** these authors contributed equally (Jawed Nawabi, Sophia Schulze-Weddige). **Corresponding Author** Sophia Schulze-Weddige Department of Radiology Universitätsmedizin Berlin Augustenburger Platz 1 13353 Berlin, Germany.

## Abstract

**Purpose:** To develop and evaluate an automated segmentation and classification tool for the discrimination of neoplastic and non-neoplastic intracerebral hemorrhage (ICH) on admission Computed Tomography (CT) utilizing images containing hemorrhage and perihematomal edema.

**Materials and Methods:** The models were developed and evaluated using a retrospective dataset of patients who presented with acute ICH of unknown cause upon admission, using CT scans obtained from a single institution between January 2016 and May 2020 for both training and testing. Etiology of ICH were binarized into non-neoplastic and neoplastic ICH according to follow-up MRI results based on the ATOMIC ICH classification. Masks for ICH and PHE were manually segmented. Two separate models were trained: 1) An nnU-Net segmentation model 2) A ResNet-34 classification model. An end-to-end tool was evaluated by concatenating the two models which allowed the segmentation model to preprocess the images for the classification model. Performance enhancement was assessed by fine-tuning the model on a randomly selected, small subset of the external cohort. To assess the model’s generalizability, the performance was additionally validated on an external dataset. Evaluation metrics were accuracy (Acc), area under the curve (AUC) and corresponding sensitivities and specificities.

**Results:** A total of 291 patients were included of whom 116 (39.86%) presented with neoplastic and 175 (60.14%) with non-neoplastic ICH. The end-to-end classification tool achieved an Acc of 86% and an AUC of 85% with a sensitivity and specificity of 80% and 93% in the test set. On the external validation cohort (n=58), the classification pipeline achieved an AUC of 68% and Acc of 66% (sensitivity 64%; specificity 67%). Fine-tuning on a selected small subset of the external cohort enhanced performance, achieving an AUC and accuracy of 70% (sensitivity 70%; specificity 71%).

**Conclusion:** An end-to-end classification tool achieved a high diagnostic performance and generalizability in classifying neoplastic from non-neoplastic ICH on CT, suggesting a robust framework for a potential clinical implementation as a decision-aided tool in early ICH management.

## 1. Introduction

Neoplastic hemorrhage is a variant of intracerebral hemorrhage (ICH) and reported to occur in up to 30% of all brain tumors, specifically in glioblastoma multiforme (GBM) and brain metastasis (BM).^1,2,3^ Diagnostic key dilemmas in these patients include stroke-like symptoms accompanied by bleedings masking the underlying tumor on imaging.^4,5,6,7,8,9,10^ Therefore, neoplastic hemorrhage may not always disclose the underlying cause of the bleeding immediately, as it may indicate, among others, a hypertensive ICH.^4^ This is of particular importance in patients with brain tumors presenting with hemorrhage as the first clinical manifestation.^4,5,6,7,8,9^ Perihematomal edema (PHE) on Computed Tomography (CT) has been introduced as a promising candidate to accurately discriminate between neoplastic and non-neoplastic hemorrhage upon admission imaging.^11,12,13^ This issue was initially explored for qualitative imaging characteristics and was further advanced into a machine learning approach adding quantitative imaging-based texture features.^11,12,13^ Recent developments of deep learning approaches have gained great utility within cancer imaging, but have not yet been explored for the discrimination of neoplastic hemorrhage.^14^ Therefore, we hypothesized that an end-to-end machine learning pipeline accurately discriminates neoplastic from non-neoplastic hemorrhage.

## 2. Material and Methods

This study was approved by the ethics committee (Charité Berlin, Germany [protocol number EA1/035/20] and written informed consent was waived by the institutional review boards. All study protocols and procedures were conducted in accordance with the Declaration of Helsinki. Patient consent was waived due to the retrospective nature of the study.

### 2.2 Study Populations

#### 2.2.1 Internal Training and Testing Cohort

Patients with acute ICH referred to Charité University Hospital Berlin, Germany were retrospectively reviewed during the period of 01/2016 to 05/2020. 27,174 patients were included according to a referral diagnosis of ICH on CT and a follow-up Magnetic Resonance Imaging (MRI). CT images were screened for the presence of parenchymal ICH and excluded when in coappearance of subarachnoid hemorrhage (SAH), subdural hematoma (SDH), or epidural hematoma (EDH). Patients with an MRI follow-up more than 30 days after initial CT imaging were also excluded. Further exclusion was performed for subjects with parenchymal ICH secondary to hemorrhagic transformation following ischemic stroke, traumatic or aneurysmal associated ICH, and cranial surgery performed prior to the admission imaging. The retrieved follow-up MRI reports were used to define the ICH etiology as previously reported^13^. In brief, ICH etiology was determined according to seven prespecified categories based on an adapted version of the mechanistic H-ATOMIC classification and described in detail in the supplementary material^15,16^. Medical records provided patient age, sex, medical history, medication details, Glasgow Come Scale (GCS) score, and symptom onset time.

#### 2.2.2 External Validation Cohort

We tested the performance and generalizability of our tool on an external data cohort, retrospectively collected from the University Medical Center Hamburg, Germany between 01/2015 to 12/2017. Inclusion criteria were consistently applied in line with the protocols established for our development cohort, ensuring uniformity across both groups.

### 2.3 Image Analysis

Non-contrast CT (NCCT) and MRI imaging data were retrieved from the local picture archiving and communication system (PACS) servers in Digital Imaging and Communications in Medicine (DICOM) format. DICOM data was anonymized in compliance with the local guidelines and transformed into Neuroimaging Informatics Technology Initiative (NifTI) files. ICH location and presence of an intraventricular hemorrhage (IVH) was documented. ICH and PHE were semi-manually quantified by planimetric measurements using the NifTI data as described in detail in the supplementary material^17,18^. A repeated segmentation was performed 3 months apart by one rater for 29 patients, selected in the test set, to calculate intra-reader agreement. Regions of interest (ROIs) were delineated using the MITK Workbench 2016.11.0 Software^19^.

### 2.4 End-to-end Pipeline

Two models were trained with manually segmented non-enhanced CT (NECT) images containing ICH and PHE regions. The first model was a segmentation model which was subsequentially used to preprocess the images for the second model, which performed the classification task. The two models are described in more detail below. The performance on the test set was evaluated by combining both models sequentially to build a fully automated end-to-end classification tool. This way of reducing complexity is a sensible strategy, particularly in the medical domain where labeled datasets are often limited in size. By applying masks to focus solely on the relevant regions of interest, we not only streamline the computational process but also optimize the utilization of available data. This targeted approach ensures that the computational burden is concentrated where it matters most, improving the efficiency of analysis and model training.

### 2.5 Preprocessing

An image subset of size 200x200x20 centered on the ICH region was created. The new images encompassed the complete ICH and PHE regions, thereby diminishing the initial input image size from 8.1 million voxels to 800,000. To achieve further reduction, we applied a mask that zeroed out all voxels beyond the boundaries of the ICH and PHE regions. As the image size of 200x200x20 was selected to accommodate even the largest lesions, most images contain significantly smaller regions of interest. Consequently, this approach served to further diminish complexity. In the training phase, the preprocessing was based on manual segmentations. In the testing phase, however, the segmentations were automatically generated by the segmentation model as discussed above.

### 2.6 Automated Deep Learning Segmentation

In the end-to-end tool a deep learning framework was employed to automatically segment the ICH and PHE region for the preprocessing of the CT images.^20^ Therefore, a MIC-DKFZ nnU-Net was trained (Division of Medical Image Computing, German Cancer Research Center (DKFZ), Heidelberg, Germany) which is a robust adaptive and open-source tool rendering state-of-the-art biomedical image segmentation^20,21^ and has already shown promising results in the context of ICH and PHE segmentation.^22^ The source code and comprehensive documentation are publicly available on GitHub.^23^ The dataset was randomly split into a training (90%) and test (10%) set as presented in figure 1. Mean and median dice score coefficients (DSC) were selected as the computational metrics for the segmentation model and computed as described in the supplementary material.^24^ DSC of the manually segmented ground truth masks were calculated for the test set of 29 subjects with non-neoplastic and neoplastic ICH.

**Figure 1:**
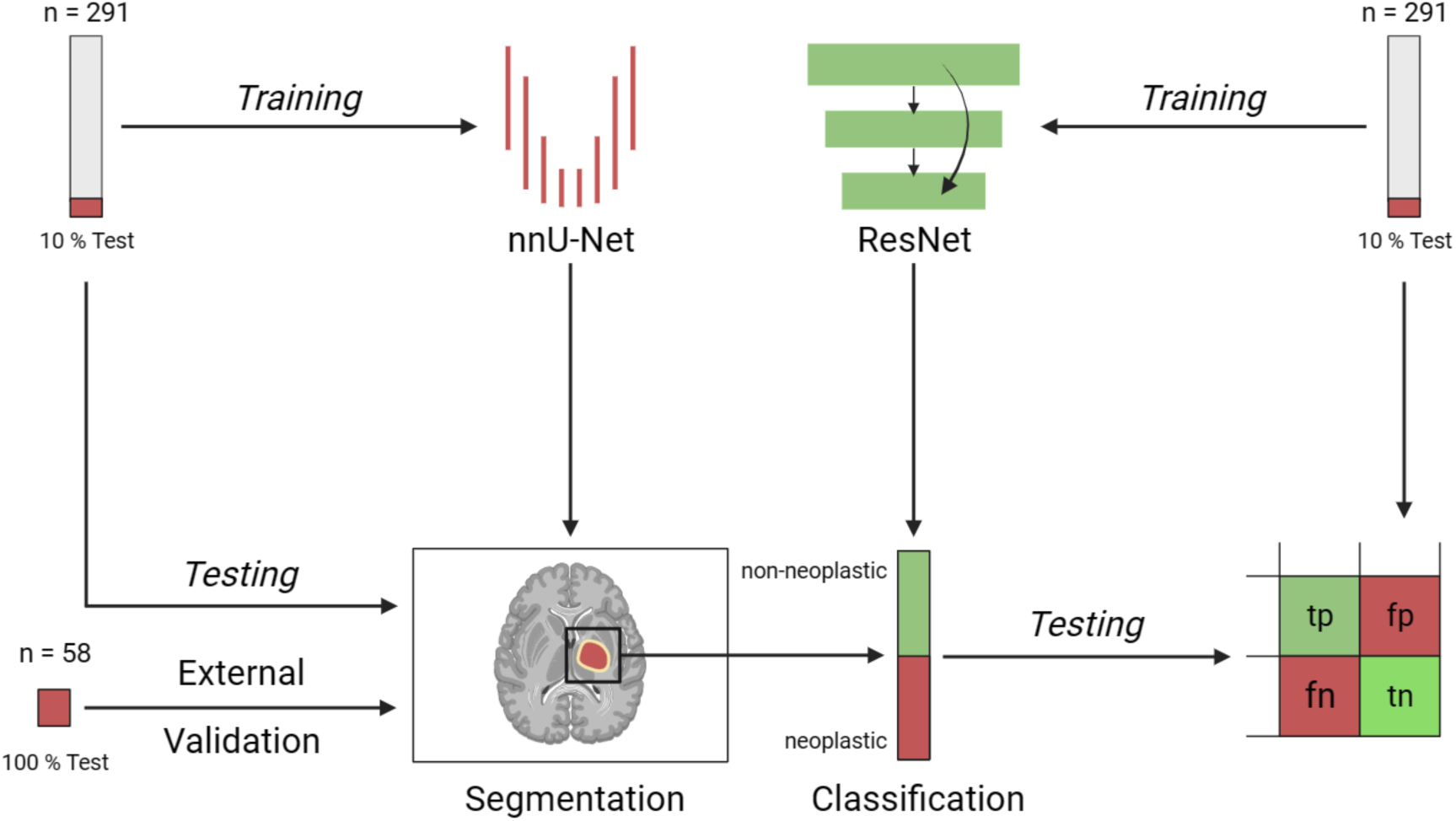
Overview of the pipeline for the classification of neoplastic and non-neoplastic and intracerebral hemorrhage. *Legend:* Pipeline for the classification of neoplastic and non-neoplastic intracerebral hemorrhage by integrating a nnU-Net-based segmentation of intracerebral hemorrhage and perihematomal edema with a deep residual network comprising 34 layers (ResNet-34) for advanced pattern recognition. The training and testing phases were conducted on a dataset comprising 291 scans, followed by an external validation on an independent dataset of 58 scans from a separate center to ensure robustness and generalizability of our model.

### 2.7 Residual Network Classification

For our study, a ResNet34 model was trained as presented in figure 2. The same training (90%) and testing (10%) split as for the segmentation model was used as presented in figure 1. Hyperparametertuning was performed to determine the best model configuration for the classification task at hand. The final model was trained for 182 epochs with a batch size of 64, a learning rate of 0.000296, the Adams optimizer, Negative Log-Likelihood loss, and image augmentations using batchgenerators^43^. As shown in figure 3, there were four indices for comparing real labels and predicted labels in the binary classification pipeline: true positive (TP), true negative (TN), false positive (FP), and false negative (FN). Performance metrics were accuracy (Acc) and area under the curve (AUC) with corresponding sensitivities and specificities. In order to optimize the balance between sensitivity and specificity on the test set and the external validation cohort, the optimal threshold was defined as the point maximizing the product of sensitivity and specificity.^25,26^

**Figure 2:**
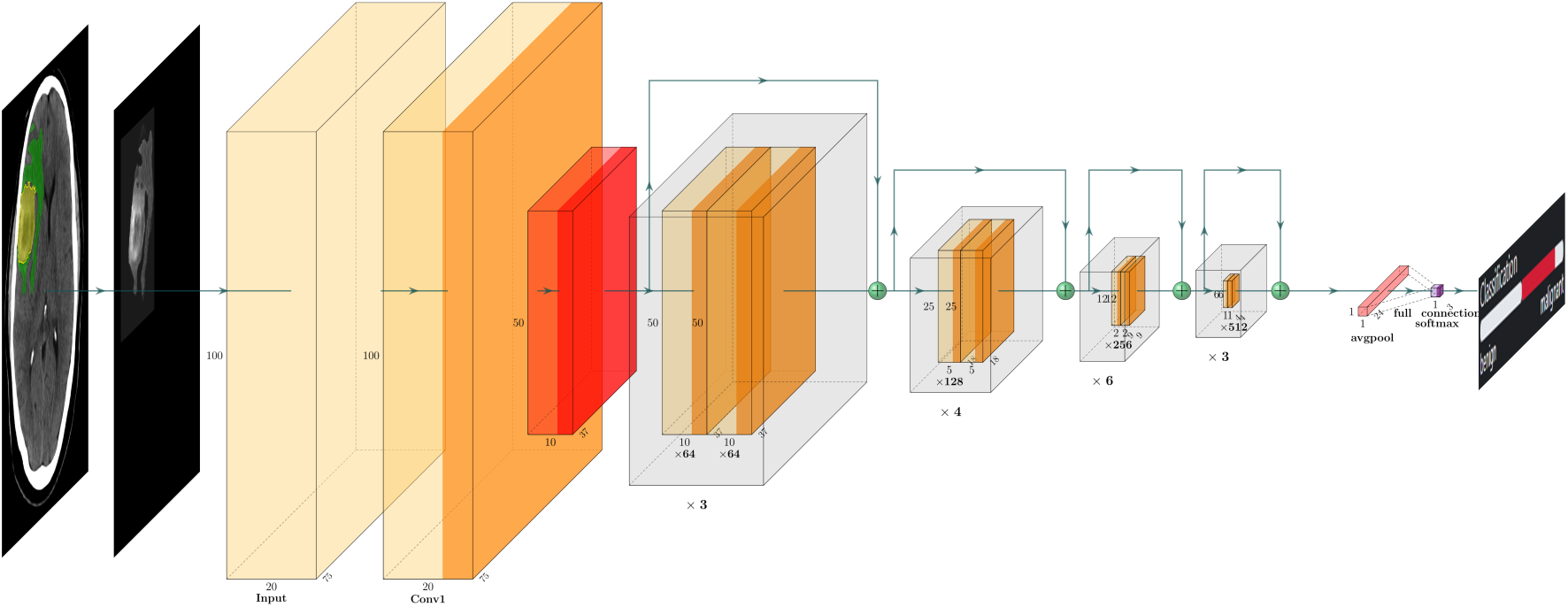
Classification model for the classification of neoplastic and non-neoplastic intracerebral hemorrhage. *Legend:* Classification of neoplastic and non-neoplastic intracerebral hemorrhage (ICH) with a deep residual network with 34 layers (ResNet-34). The graphic illustrates the ResNet34 model architecture. The input layer processes segmented medical images containing only the ICH and PHE region, followed by convolutional blocks with residual connections for effective feature extraction. Pooling layers (red) reduce spatial dimensions, and fully connected layers make the classification decision. The output layer uses softmax activation for probability predictions of neoplastic and non-neoplastic classes. The prediction is displayed on a scale from benign to malignant in our web based custom reader interface (figure 4).

**Figure 3:**
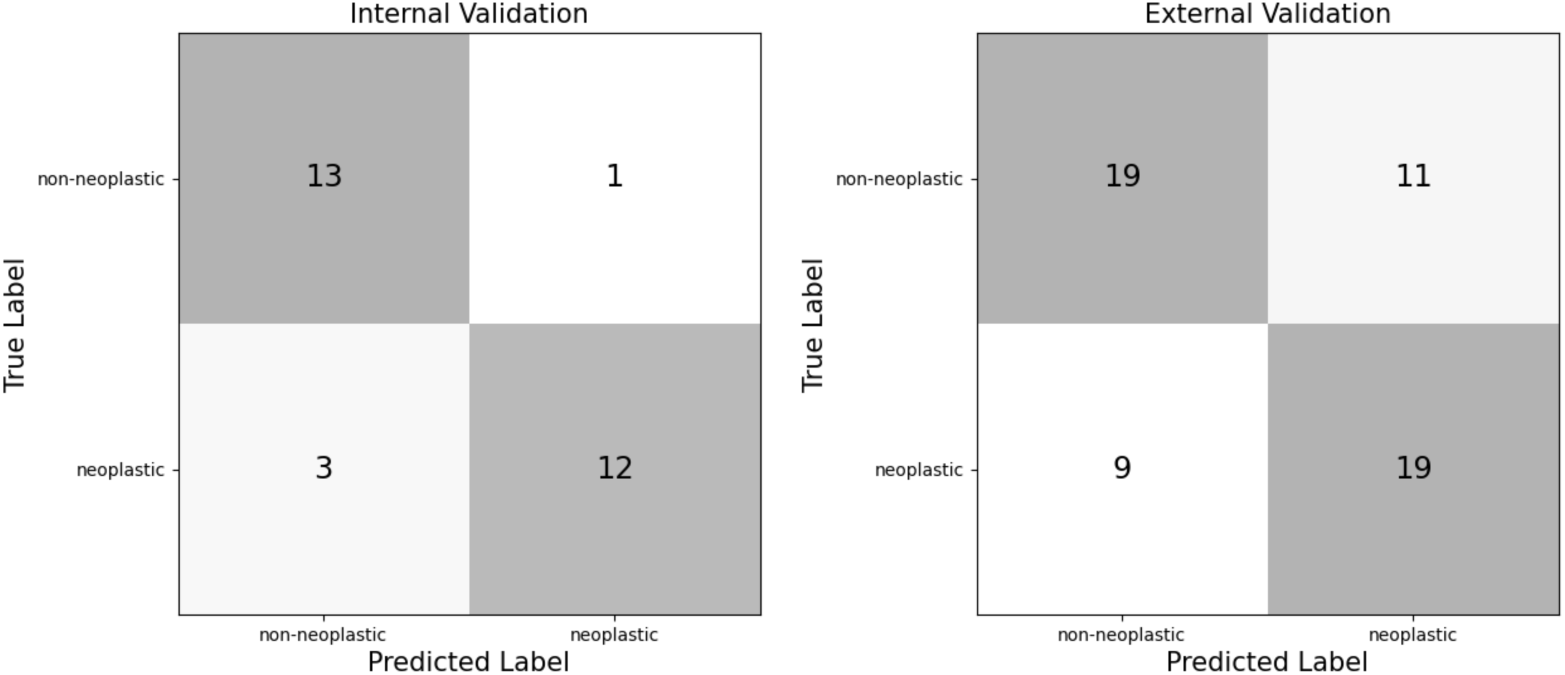
Confusion matrices for classification of non-neoplastic and neoplastic intracerebral hemorrhage. *Legend:* Confusion matrices for the classifier in the test set (left) and the external validation cohort (right).

### 2.8 Statistical Analysis

For the clinical indicators of patients, data was tested for normality and homogeneity of variance using histogram plots and the Shapiro-Wilk test. Descriptive statistics are presented as counts (percentages [%]) for categorical variables and compared with χ^2^ test. Continuous variables are presented as mean (standard deviation [SD]) for normally distributed variables, and medians (interquartile range [IQR]) for non-normal variables which are compared with the Mann-Whitney test. A *p-*value < 0.05 was considered as significant. Statistical analyses were performed using the IBM SPSS Statistics 21 software package (IBM Corporation, Armonk, NY) and python package sklearn 0.0.post1. Visualizing the data was done in an Ubuntu 20.04.5 LTS environment using jupyterlab 3.4.5 and LaTeX 2.0 as well as the following python packages: matplotlib 3.5.2, seaborn 0.11.2. The following packages were used in the ResNet training process: torch 1.12.1+cu113, batchgenerators 0.24, scikit-learn 1.1.2, NumPy 1.23.3, pandas 1.5.0. The following packages were used in the nnU-Net training process: nnU-Net Version 1.6.6, SimpleITK Version 2.0.2, pandas v 1.2.3, NumPy 1.20.2.

## 3. Results

### 3.1 Patients Characteristics of the Internal Training and External Validation Cohort

Characteristics of 291 patients in training and testing cohorts are in Table 1. 116 (39.86%) had neoplastic ICH, predominantly brain metastases (79; 68.10%) versus primary tumors (37; 31.90%). Non-neoplastic ICH was found in 175 (60.14%). Neoplastic ICH had larger PHE volumes (median 23.11 ml, IQR: 12.45-60.25 ml) than non-neoplastic (median 10.48 ml, IQR: 5.32-24.37 ml, p < 0.001). Hematoma volumes were similar between groups (p = 0.56). The external validation cohort mirrored these findings.

**Table 1:**
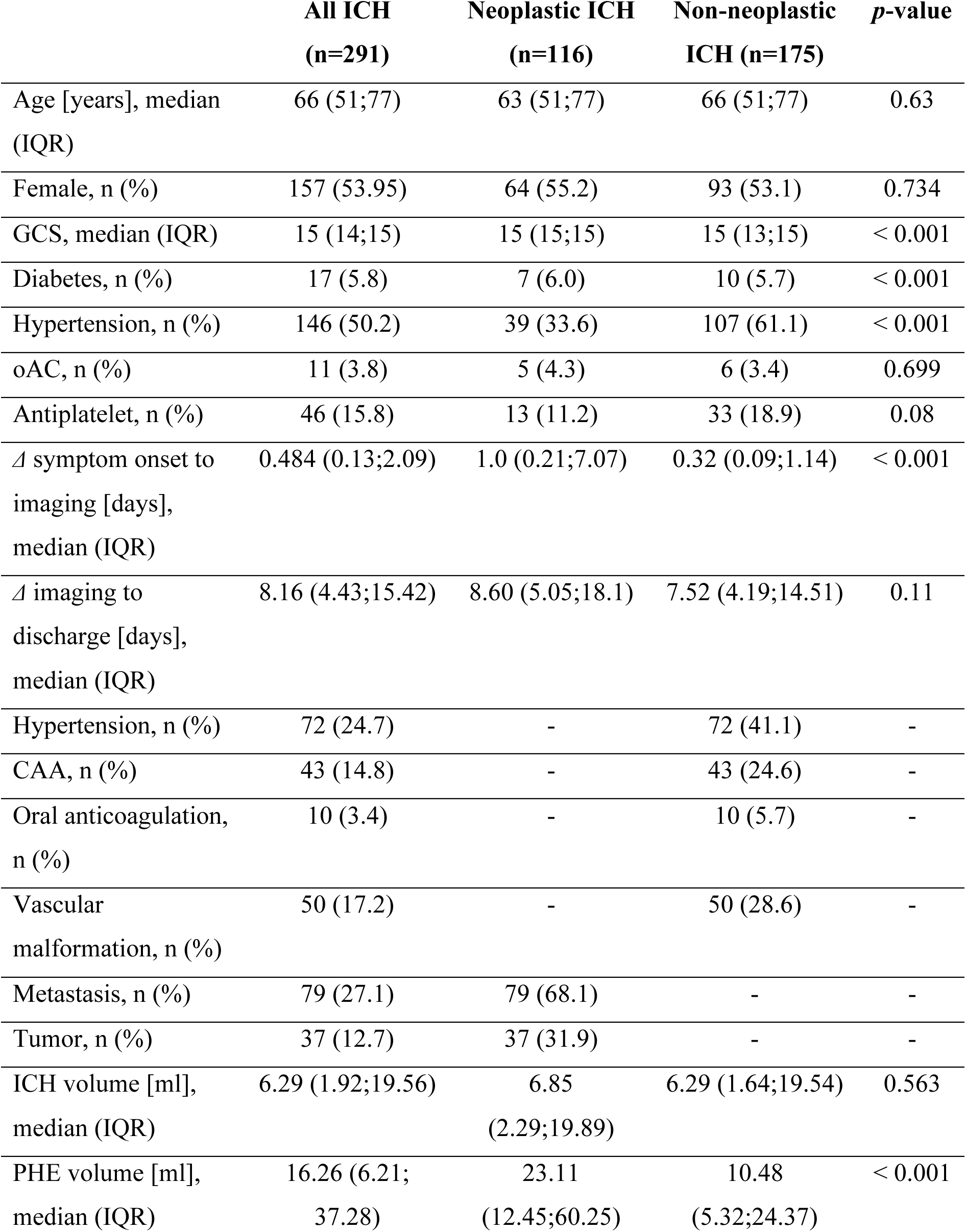
Baseline characteristics of patients with acute neoplastic and non-neoplastic intracerebral hemorrhage. *Legend:* CAA, cerebral amyloid angiopathy; *Δ,* delta; ICH, intracerebral hemorrhage; IQR; interquartile range; GCS, Glasgow Come Scale; ml, milliliters; oAC, oral anticoagulation; PHE, perihematomal edema.

|  | **All ICH**<br>**(n=291)** | **Neoplastic ICH**<br>**(n=116)** | **Non-neoplastic**<br>**ICH (n=175)** | **p-value** |
| --- | --- | --- | --- | --- |
| Age [years], median (IQR) | 66 (51;77) | 63 (51;77) | 66 (51;77) | 0.63 |
| Female, n (%) | 157 (53.95) | 64 (55.2) | 93 (53.1) | 0.734 |
| GCS, median (IQR) | 15 (14;15) | 15 (15;15) | 15 (13;15) | < 0.001 |
| Diabetes, n (%) | 17 (5.8) | 7 (6.0) | 10 (5.7) | < 0.001 |
| Hypertension, n (%) | 146 (50.2) | 39 (33.6) | 107 (61.1) | < 0.001 |
| oAC, n (%) | 11 (3.8) | 5 (4.3) | 6 (3.4) | 0.699 |
| Antiplatelet, n (%) | 46 (15.8) | 13 (11.2) | 33 (18.9) | 0.08 |
| $\Delta$ symptom onset to imaging [days], median (IQR) | 0.484 (0.13;2.09) | 1.0 (0.21;7.07) | 0.32 (0.09;1.14) | < 0.001 |
| $\Delta$ imaging to discharge [days], median (IQR) | 8.16 (4.43;15.42) | 8.60 (5.05;18.1) | 7.52 (4.19;14.51) | 0.11 |
| Hypertension, n (%) | 72 (24.7) | - | 72 (41.1) | - |
| CAA, n (%) | 43 (14.8) | - | 43 (24.6) | - |
| Oral anticoagulation, n (%) | 10 (3.4) | - | 10 (5.7) | - |
| Vascular malformation, n (%) | 50 (17.2) | - | 50 (28.6) | - |
| Metastasis, n (%) | 79 (27.1) | 79 (68.1) | - | - |
| Tumor, n (%) | 37 (12.7) | 37 (31.9) | - | - |
| ICH volume [ml], median (IQR) | 6.29 (1.92;19.56) | 6.85 (2.29;19.89) | 6.29 (1.64;19.54) | 0.563 |
| PHE volume [ml], median (IQR) | 16.26 (6.21; 37.28) | 23.11 (12.45;60.25) | 10.48 (5.32;24.37) | < 0.001 |

### 3.2 Automated Lesion Segmentation of ICH and PHE

On the test set, the segmentation model achieved median DSC of 0.80 (mean 0.70 ± 0.30; range 0 – 0.95) and 0.70 (mean 0.60 ± 0.24; range 0 – 0.86) for ICH and PHE, respectively. Median DSC of ground truth masks for repeated segmentations were 0.86 (mean 0.85 ± 0.11; range 0.43 – 0.96) for ICH and 0.71 (mean 0.69 ± 0.12; range 0.36 – 0.85) for PHE.

On the external validation cohort, the model achieved median DSC of 0.71 (mean 0.66 ± 0.20; range 0 – 0.88) and 0.60 (mean 0.55±0.19; range 0 – 0.87) for ICH and PHE, respectively. In two cases, the model did not segment any ICH or PHE region. These cases were excluded from the proceeding analysis of the classification tool due to the absence of segmented regions in the preprocessed images.

### 3.3 End-to-end Tool for the Classification of Neoplastic and Non-neoplastic ICH

Our end-to-end tool distinguished neoplastic from non-neoplastic ICH with an AUC of 85% and Acc of 86% (sensitivity 80%; specificity 93%) in the test set with a decision threshold of - 1.5 on the model’s output of the positive class. On the external validation cohort, the classification pipeline achieved an AUC of 68% and Acc of 66% (sensitivity 68%; specificity 63%). These results were reached with a decision threshold of -0.075. The end-to-end classification tool failed to correctly classify four subjects which have been analyzed in detail and presented on a case-by-case basis in the supplementary figures 1-4. It misclassified one subject as neoplastic ICH (false positive; FP) and three as non-neoplastic ICH (false negative; FN). Resulting performance metrics are shown in table 2. The results for the indices of the confusion matrix are presented in figure 3. An illustrative example of the web based custom reader interface for the automated segmentation and classification of neoplastic and non-neoplastic ICH is illustrated in figure 4.

**Figure 4:**
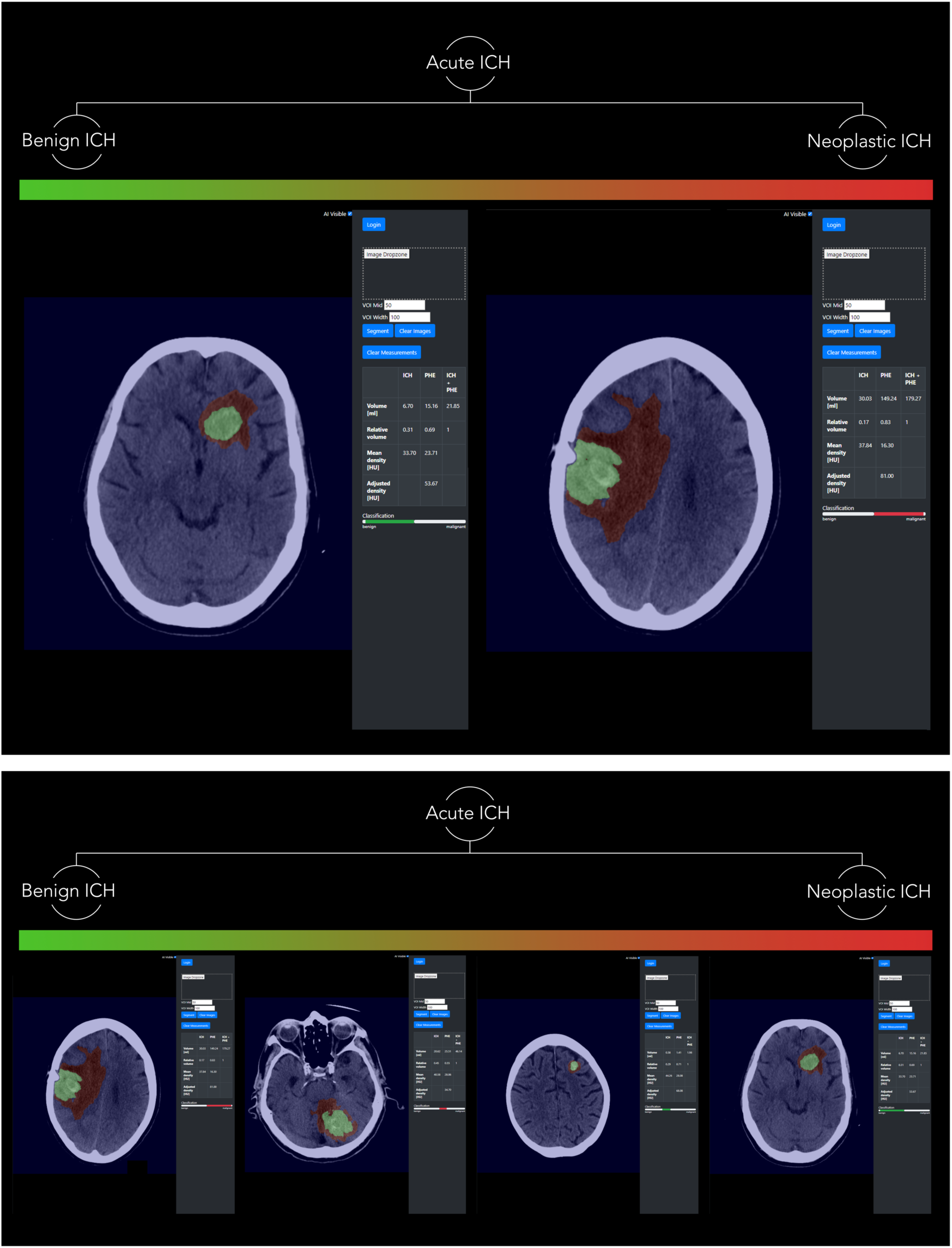
Web based custom reader interface for the fully automated prediction of neoplastic and non-neoplastic intracerebral hemorrhage on Computed Tomography. *Legend:* Imaging panel of web based custom reader interface with an image drop zone for the automated lesion detection, segmentation, and calculation of parenchymal hemorrhage (green) and perihematomal edema (red) volumes and densities - displayed to the reader on the right imaging panel. Automated prediction of non-neoplastic (green coded bar displayed below the volume characteristics; left) and neoplastic bleeding (red coded bar displayed below the volume characteristics; right) with a high certainty (upper panel) compared to two additional cases of less certainty (lower panel). ICH; intracerebral hemorrhage; HU, Hounsfield Unit; PHE, perihematomal edema

**Table 2:**
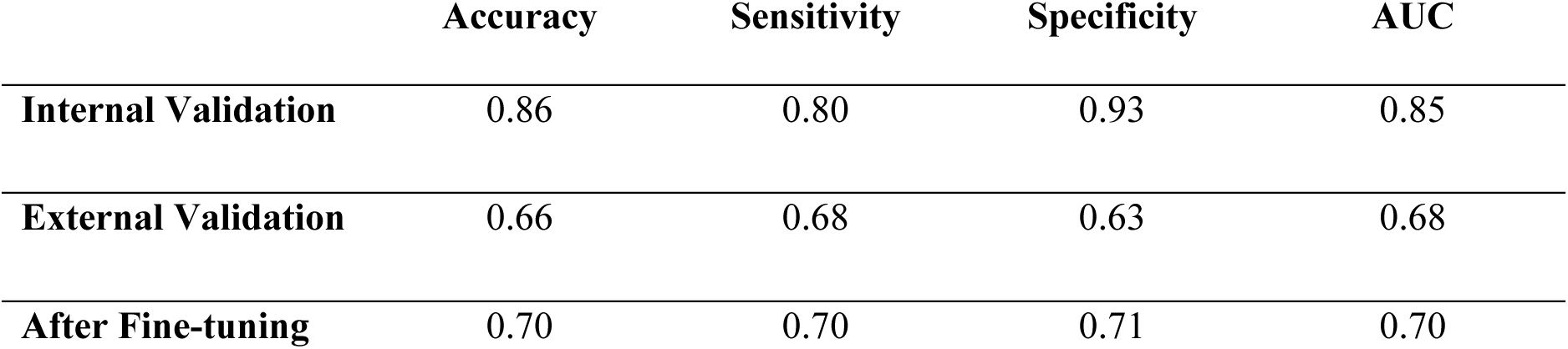
Diagnostic performance of the end-to-end prediction model in the classification of neoplastic and non-neoplastic intracerebral hemorrhage in the internal test dataset and the external validation cohort. *Legend:* AUC; area under the curve.

## 4. Discussion

In this study, we proposed a prediction model for the automated segmentation and classification of neoplastic ICH utilizing imaging properties on CT from the parenchymal hemorrhage and surrounding edema formation in an ICH patient cohort. The proposed prediction pipeline demonstrated an overall high accuracy and diagnostic performance, suggesting a robust framework for an end-to-end tool to be implemented in the clinic. As shown in the confusion matrices, only a small number of subjects were incorrectly classified which have been compiled on a case-by-case basis in the supplementary figures. In the test set, one single subject with non-neoplastic ICH was incorrectly classified. The corresponding CT image displayed a bleeding in the paravermal cerebellar hemisphere which was confirmed as a rare pial-type of an arteriovenous malformation as indicated in the supplementary figure 1. On CT imaging the bleeding was surrounded by relatively pronounced PHE formation which potentially could be a result of a repeated bleeding event found in arteriovenous malformations.^27^

Our previous results have confirmed higher PHE characteristics in neoplastic ICH to be of high discriminatory value in the classification of neoplastic and non-neoplastic ICH and therefore may have equally led to the misclassification of the current classifier.^11,17,13^ Among the falsely negative classified subjects, two were characterized by small bleedings of metastatic ICH as presented in the supplementary figures 2 and 3. As the preprocessing exclusively segmented the largest lesion, the classification model generated predictions based solely on this particular instance of bleeding rather than considering the entirety of the bleeding instances. This complicated the classification task, potentially contributing to the misclassification errors. Another case of metastases associated ICH was incorrectly classified as presented in the supplementary figure 4. This patient with an unknown melanoma metastasis presented with a large parenchymal bleeding and extension to the ventricular system - both displaying very atypical imaging findings of an underlying neoplastic brain lesion and being rather indicative, among others, of a hypertensive or amyloid angiopathy associated bleeding.^29^ In hindsight, the patient’s record indicated that the patient had concomitantly suffered from meningitis ventriculitis which was unknown at the point of the follow-up imaging that we used for our classification scheme. It is to be believed that these complications may have caused or at least have strongly influenced the development of a large parenchymal ICH with extension to the ventricular system.^30,31^ In compliance with the aforementioned, IVH among patients with neoplastic brain lesions is limited to two case reports. In a case series of 15 patients with metastatic bleedings, five patients presented with some of the parenchymal hemorrhage ruptured into the ventricular system, and one presented with a primary IVH only.^10^ The authors found no presence of IVH among a subgroup of patients with glioma associated bleedings.^10^ In line with this, IVH among primary brain tumors is only reported in one case report of intraventricular meningioma.^32^ A rare condition seen in 0.5–5% of all intracranial meningiomas with 80% presenting in the lateral ventricles - similarly to our case.^32^ Hence, massive IVH is a very random condition in neoplastic brain lesions and may result primarily from a metastatic tumor at or within the cerebral ventricles. Considering the rather rarely seen and clinically inconclusive cases, coupled with small bleedings primarily in metastatic lesions, it is conceivable that our proposed classification tool showed an overall good performance.

The widespread adoption of ML tools in medical imaging, particularly for the analysis of CT images, is hindered by the challenge of poor generalizability across different scanners and local clinical practices^44^. The inherent variability in imaging protocols, hardware configurations, and acquisition techniques among diverse medical institutions leads to a lack of standardization in the data. ML models trained on images from one scanner or clinic may struggle to adapt effectively to data from others, compromising their performance and reliability. This issue is exacerbated by subtle yet consequential differences in image quality, resolution, and noise levels between machines^45^. As a result, the robustness and accuracy demonstrated during model development may not translate seamlessly when applied to CT images acquired from disparate sources which also translated into a reduced performance on our external dataset. While a 17% reduction in AUC and Acc is not optimal, the performance in the external validation remains satisfactory, given the diverse data sources. To enhance performance, we attempted model finetuning using a limited subset of images from the external dataset. Following a common approach, we froze the trained layers of the classification model and selectively trained the last layer. Specifically, we randomly selected 20% (n=12) of cases from the external cohort for finetuning, resulting in a 2% improvement in AUC. Further enhancements are anticipated with more training data. We deliberately refrained from utilizing more external data to maintain a realistic evaluation of performance.

This study has several strengths. First, we utilized two state-of-the-art deep convolutional neural networks. The ResNet was developed with the main intent of designing very-deep networks that did not suffer from the “vanishing gradient” problem that was prevalent in its predecessors.^33^ The nnU-Net outperformed most deep learning networks and won several competitions of segmentation tasks of biomedical imaging.^21^ In addition, we added imaging information on PHE which have shown to be important in the differential diagnosis of neoplastic brain lesions. ^11,17^ In our previous studies the diagnostic properties of PHE had been initially analyzed in a conventional and later radiomics based machine learning approach. ^11,17^ Despite the promising results both methods lacked the potential for a clinical implementation as the manual lesion segmentation was a prerequisite for the classification task.^12^ Previous studies had proposed automated algorithms for the segmentation and volumetric estimation of both ICH and PHE, but their methods were limited to non-neoplastic, and foremost hypertensive, ICH patients.^32,40,41^ Hence, the performance in a real-world ICH patient cohort, especially with neoplastic ICH, was supposedly uncertain. To our knowledge this is the first study to propose a fully automated tool for the classification of neoplastic and non-neoplastic ICH. This can be considered as a significant step forward in improving the diagnostic dilemma and hence clinical management and outcome of patients with neoplastic brain lesions presenting with ICH.^4,5,6,7,8,9^ In line with this, we implemented a ready to use web-based custom reader interface to be integrated in the clinical workflow and allow a quick and easy imaging interpretation in the real-life clinical setting. We plan to improve the model by adding subgroup specific classifications for all ICH etiologies. This is of clinical interest, as the differentiation of metastatic and primary brain tumors is important since clinical management and treatment of these two types of tumors are radically different.^37,43^ The same applies for non-neoplastic ICH which represent a very large group of heterogenous entities and could help to inform treating physicians about prognosis and treatment strategies.^16,42^ Another strength is that the generalizability of our approach was tested in an external cohort. This step is often omitted which means no claims about the general performance of a given model can be made. Although the performance dropped, we are still content with the results as an equally high performance on images from different scanners is unlikely in most AI models. Further, fine-tuning on as little as 12 cases already improved the performance. Hence, with limited effort the approach can be translated to other datasets, clinics, and scanners.

Our study had some limitations. Firstly, as this was a clinical study of retrospectively collected data it bears a risk of bias. ICH etiology was determined by follow-up MRI report. Additional pathology reports may have increased the certainty of ICH etiology but were not universally available. Secondly, the DSC showed potential for improvement in the PHE segmentation quality. However, these results agree well, and in some cases even prove better, with previous results reporting fully automated PHE segmentation models.^22,38,39^ As stated above, the proposed model performed less in cases of small ICH, primarily with metastatic lesions as presented in the supplementary figure 2 and 3, a problem that has been frequently been found in deep learning segmentation models.^22^ This issue should be addressed in future studies by adding more, heterogenous training data to further enhance the model performance.

In conclusion, our fully automated approach demonstrated a high performance in the classification of neoplastic and non-neoplastic ICH suggesting a robust end-to-end framework for the integration in the clinical workflow as a decision-aided tool via our proposed user-friendly web-based custom reader interface. In order to be applied in different clinics, fine-tuning on a small set of samples is recommended to adapt to the specificities of the new dataset such as scanner type and protocol.

## Data Availability

The datasets that support the findings of our study are available upon reasonable request from the corresponding author.

## Authorship

All authors contributed to the study conception and design. Material preparation, data collection and analysis were performed by Tobias Orth, Jawed Nawabi, Sophia Schulze-Weddige, Tobias Penzkofer and Geor g Lukas Baumgaertner. The first draft of the manuscript was written by Jawed Nawabi and Sophia Schulze-Weddige and all authors commented on previous versions of the manuscript. All authors read and approved the final manuscript.

## Abbreviations

Acc: accuracy
AUC: area under the curve
BM: brain metastasis
CAA: cerebral amyloid angiopathy
CNN: convolutional neural network
CT: Computed Tomography
DKFZ: Division of Medical Image Computing, German Cancer Research Center
DSC: dice score coefficients
EDH: epidural hematoma
ICH: intracerebral hemorrhage
IQR: interquartile range
IVH: intraventricular hemorrhage
FN: false negative
FP: false positive
GBM: glioblastoma multiforme
GCS: Glasgow Come Scale
HU: Hounsfield units
MRI: Magnetic Resonance Imaging
NCCT: non-contrast CT
NCET: non-enhanced CT
NifTI: Neuroimaging Informatics Technology Initiative
PACS: picture archiving and communication system
PHE: perihematomal edema
ResNet: Deep Residual Network
ROI: region of interest
SAH: subarachnoid hemorrhage
SDH: subdural hematoma
SD: standard deviation
TN: true negative
TP: true positive

