## Supplementary_Material for "End-to-End Machine Learning based Discrimination of Neoplastic and Non-neoplastic Intracerebral Hemorrhage on Computed Tomography"

#### Study population

For the purpose of this study, we did not mandate a strict hierarchy as in the original H-ATOMIC classification but the etiology, which was most likely causal based on evidence from the follow-up MRI exam.[16] Subjects with non-neoplastic ICH were classified as (1) hypertensive ICH, (2) cerebral amyloid angiopathy (CAA)-, (3) oral anticoagulation-, or (4) vascular malformation associated ICH. Subjects with neoplastic ICH were classified as either (5) metastatic or (6) primary brain tumor associated ICH. Subjects without a possible, probable, or definite etiology of ICH were referred to as crypotogenic (7). Patients with cryptogenic and unconfirmed ICH on MRI were excluded from the study as previously described[44].

#### Image Analysis

The dataset was inspected for quality and excluded in case of severe motion artifacts, post-contrast CT, inadequate slice thickness (> 5 mm thickness), extra-axial brain tumor or cranial surgery apparent on admission CT. The ROI histogram for PHE segmentation was sampled between 0 and 30 Hounsfield units (HU) to exclude voxels that likely belong to leucariosis[17]. The ROI histogram for ICH segmentation was sampled between 20 and 80 HU to exclude voxels that likely belonged to cerebrospinal fluid or calcification[18]. Imaging analysis was performed by a research student (TO with 3 years of experience in ICH imaging research) and inspected for correction by a radiology fellow (JN with 6 years of expertise neuroimaging and ICH imaging research). Both readers were blinded to all clinical information and MRI scans.

#### Automated Deep Learning Segmentation

DSC was calculated as the following:

$$Dice\ Score = \frac{2\,(\mathrm{P} \cap \mathrm{T})}{(|\mathrm{P}| + |\mathrm{T}|)}$$

Where ∩ is the intersection and *(P ∩ T)* represents the spatial overlap between *P* and *T*. *T* denotes the ground truth segmentation and *P* denotes the predicted segmentation. $|P|$ and $|T|$ represent the areas of *P* and T, respectively. For the end-to-end approach, a web based custom reader interface was developed to display the segmentations of ICH and PHE alongside the volumetric estimation and classification prediction for the prediction of a neoplastic ICH.

### Supplementary Tables and Figures

**Supplementary Table 1:** Baseline characteristics of the external validation cohort.

| | **All ICH (n=58)** | **Neoplastic ICH (n=28)** | **Non-neoplastic ICH (n=30)** | ***p*-value** |
|---|---|---|---|---|
| Age [years], median (IQR) | 73 (55;82) | 70 (58;80) | 75 (55;83.75) | 0.102 |
| Female, n (%) | 25 (43.1) | 11 (39.3) | 33 (56.9) | 0.052 |
| *Δ* symptom onset to imaging [hours], median (IQR) | 10.50 (3;27.5) | 20.0 (4;48) | 8 (2.13;24) | 0.053 |
| Hypertension, n (%) | 11 (19) | - | 11 (36.7) | - |
| CAA, n (%) | 6 (10.3) | - | 6 (20.0) | - |
| Vascular malformation, n (%) | 13 (22.4) | - | 13 (43.4) | - |
| Metastasis, n (%) | 23 (39.7) | 23 (82.1) | - | - |
| Tumor, n (%) | 5 (8.6) | 5 (17.9) | - | - |
| ICH volume [ml], median (IQR) | 14.97 (5.69;32.05) | 15.48 (8.91;36.11) | 10.8 (5.23;31.18) | 0.641 |
| PHE volume [ml], median (IQR) | 20.35 (8.86;51.51) | 35.58 (18.99;76.89) | 12.33 (3.45;31.64) | 0.001 |

*Legend:* CAA, cerebral amyloid angiopathy; *Δ,* delta; ICH, intracerebral hemorrhage; IQR; interquartile range; GCS, Glasgow Come Scale; ml, milliliters; PHE, perihematomal edema.

**Supplementary Figure 1:** Case-by-case analysis of the few misclassified cases 1 of 4

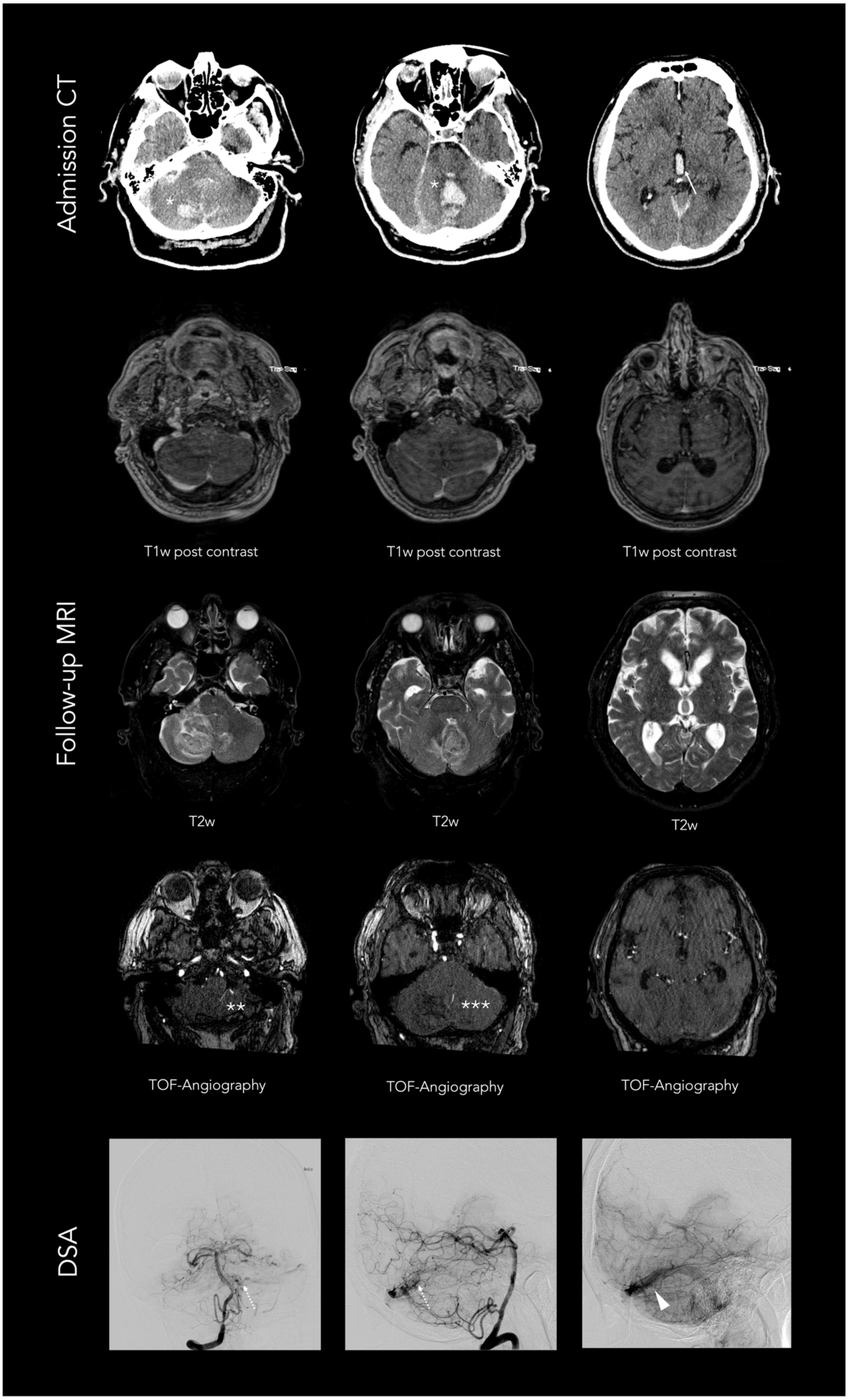

*Legend:* Non-neoplastic intracerebral hemorrhage (ICH) incorrectly classified as neoplastic ICH (false positive). ICH presenting with a cerebral parenchymal bleeding marked as star and hemorrhage extension into both lateral ventricles (marked as arrow) in anatomical order on admission Computed Tomography (CT; first panel) and on follow-up Magnetic Resonance Imaging (MRI) with, T1-weighted post contrast (second panel), T2-weighted (third panel), and time-of-flight angiography (fourth panel) sequences. Where a pial arteriovenous fistula was diagnosed with arterial supply from both the posterior inferior cerebellar artery (marked as two stars) and superior cerebellar artery (marked as three stars) and confirmation on digital subtraction angiography (dotted arrow) with drainage into the left sided transverse sinus (short arrow). DSA indicates digital subtraction angiography; T1w indicates T1-weighted imaging; T2w, T2-weighted imaging; TOF, time-of-flight.

**Supplementary Figure 2:** Case-by-case analysis of the few misclassified cases 2 of 4

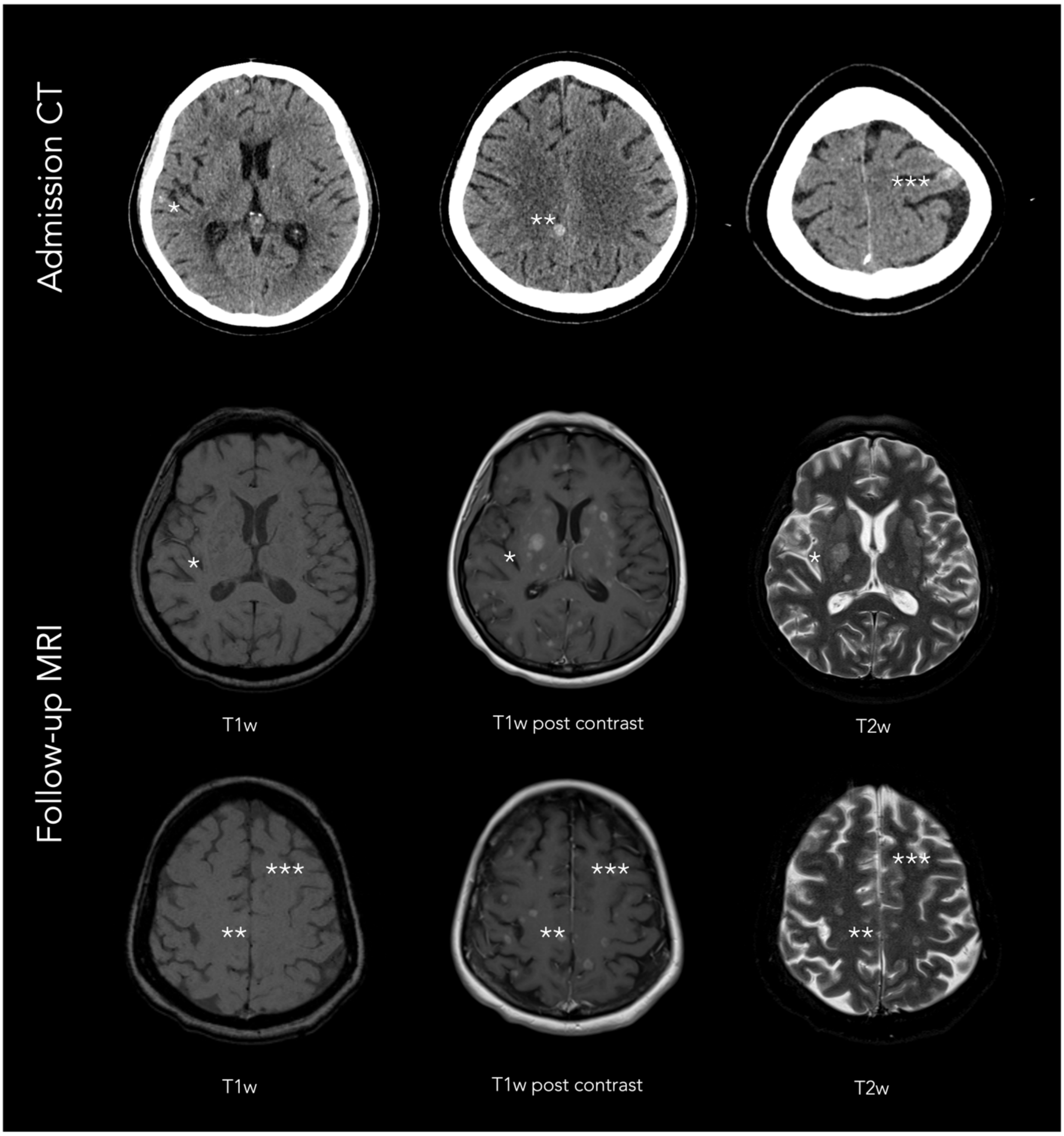


*Legend:* Tumorous intracerebral hemorrhage (ICH) incorrectly classified as non-tumorous ICH (false negative). ICH presenting with three small parenchymal bleedings marked as stars in anatomical order on admission Computed Tomography (CT; upper panel) and on follow-up Magnetic Resonance Imaging (MRI) with T1-weighted (left lower panels), T1-weighted post contrast (middle lower panels), and T2-weighted (right lower panels) sequences. Where first bleeding is located in the cortical region of the right temporal lobe (*). Second bleeding in the right parasagittal parietal lobe (**). Third bleeding in the left frontal lobe (***). T1w indicates T1-weighted imaging; T2w, T2-weighted imaging.

**Supplementary Figure 3:** Case-by-case analysis of the few misclassified cases 3 of 4

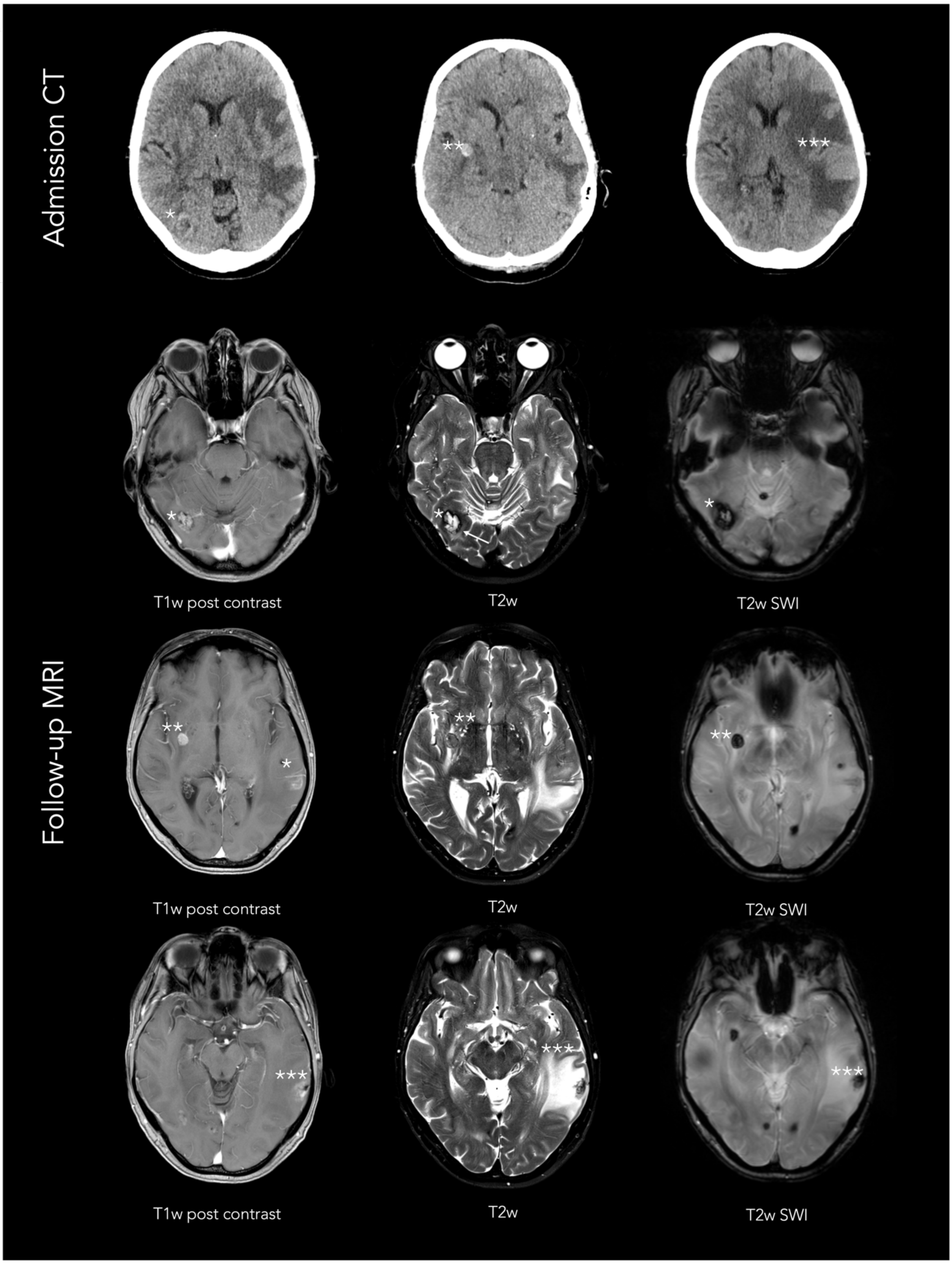


*Legend:* Tumorous intracerebral hemorrhage (ICH) incorrectly classified as non-tumorous ICH (false negative). ICH presenting with three parenchymal bleedings marked as stars in anatomical order on admission Computed Tomography (CT; upper panel) and on follow-up Magnetic Resonance Imaging (MRI) with T1-weighted post contrast (left lower panels), T2-weighted (middle lower panels), and T2-weighted susceptibly (right lower panels) sequences.

Where first bleeding is located in the right temporooccipital lobe (*). Second bleeding in the right basal ganglia (**). Third bleeding in the left temporal with a comparatively larger perihematomal and peritumoral edema formation (***). T1w indicates T1-weighted imaging; T2w, T2-weighted imaging; SWI, susceptibly weighted imaging.

**Supplementary Figure 4:** Case-by-case analysis of the few misclassified cases 4 of 4

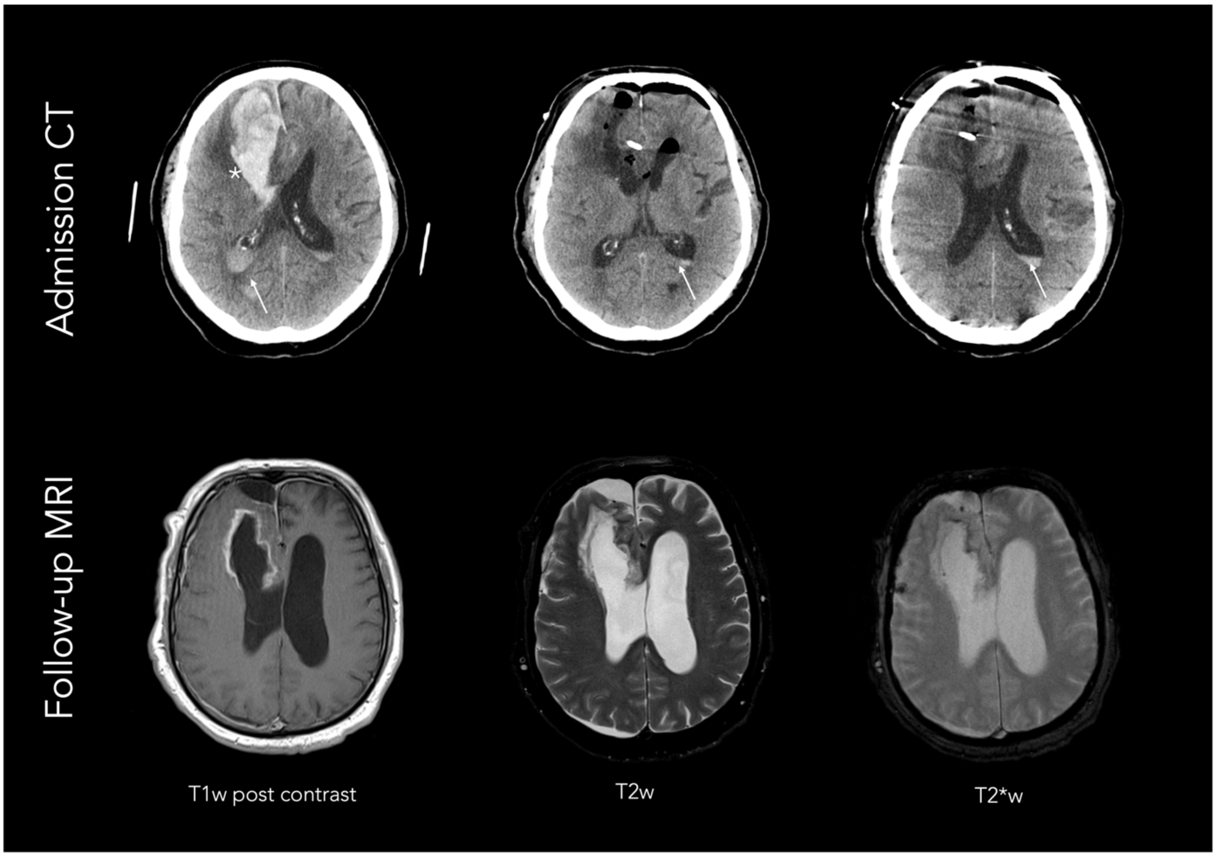


*Legend:* Tumorous intracerebral hemorrhage (ICH) incorrectly classified as non-tumorous ICH (false negative). ICH presenting with one large parenchymal bleeding (marked as star) and hemorrhage extension into both lateral ventricles (marked as arrows) on admission Computed Tomography before and after immediate surgical evacuation (CT; upper panel) as well as follow-up Magnetic Resonance Imaging (MRI) after three days of surgery with T1-weighted post contrast (left lower panel), T2-weighted (middle lower panel), and T2-weigted susceptibly (right lower panel) sequences. T1w indicates T1-weighted imaging; T2w, T2-weighted imaging; T2*w, T2*-weighted imaging.
